# A retrospective study of the clinical characteristics of COVID-19 infection in 26 children

**DOI:** 10.1101/2020.03.08.20029710

**Authors:** Anjue Tang, Wenhui Xu, Min Shen, Peifen Chen, Guobao Li, Yingxia Liu, Lei Liu

## Abstract

**Background:** The outbreak of novel coronavirus pneumonia in China began in December 2019. Studies on novel coronavirus disease (COVID-19) were less based on pediatric patients. This study aimed to reveal the clinical characteristics of COVID-19 in children.

**Method:** This study retrospectively analyzed the clinical symptoms, laboratory results, chest CT, and treatment of children with laboratory-confirmed COVID-19(ie, with samples that were positive for 2019 novel coronavirus[2019-nCoV]) who were admitted to Shenzhen Center of National Infectious Disease Clinical Medical Research from January 16 to February 8, 2020.

**Result:** Nine patients had no obvious clinical symptom. 11 patients developed fever. Other symptoms, including cough(in eleven of seventeen patients), rhinorrhea(in two), diarrhea(in two), vomiting(in two), were also observed. A small minority of patients had lymphocytopenia. Alanine transaminase or transaminase increased in three cases. According to chest CT scan, 11 patients showed unilateral pneumonia, 8 patients had no pulmonary infiltration. No serious complications such as acute respiratory syndrome and acute lung injury occurred in all patients.

**Conclusion:** The clinical characteristics of 2019-nCoV infection in children were different from adult. The overall condition of children were mild and have a good prognosis.

**Mainpoint:** COVID-19 is a kind of new infectious disease.The clinical characteristics of 2019-nCoV infection in children may different from adult. Myocardium likely less affected by 2019-nCoV in children.

## BACKGROUND

Pneumonia caused by the novel coronavirus was discovered in Wuhan, Hubei, China and then the epidemic broke out. It was defined by the World Health Organization as “an international public health emergency of public concern” [1, 2]. Recently, many researches in the lancet reported the epidemiological, clinical symptoms, laboratory data, imaging features, treatment methods and clinical outcomes of non-pregnant adults and pregnant women infected with 2019-nCOV[3, 4]. However, the clinical characteristics of children have not been well disclosed, and whether there are differences in treatment methods has not been determined. We retrospectively analyzed the clinical data of 28 children with COVID-19 tested positive for 2019-nCOV by use of RT-PCR on samples.

## METHOD

### Patient information

We reviewed 26 cases of children(>1 year old and <14 years old) in the Third People’s Hospital of Shenzhen from January 16 to February 8, 2020. Diagnosis of COVID-19 was based on the Novel Coronavirus Pneumonia Prevention and Control Program(6th edition) issued by the National Health and Health Committee of China[5]. The study was approved by the instututional of research ehtics committee of the Third People’s Hospital of Shenzhen ([2020-063]).

### Data collection

We collected the clinical manifestations, laboratory results, chest CT, treatment methods, and outcomes of 26 children, and the data was updated until February 19. To ensure the accuracy of the data, the data were independently checked by two clinicians. Each patient signed a written informed consent. We collected respiratory, blood, and digestive tract specimens from children for examination. All samples were processed simultaneously at the department of Shenzhen Center for Disease Control and Prevention(CDC) and the hepatopathy institute of National Infectious Disease Clinical Medical Research Center(The Third People’s Hospital of Shenzhen).

### Statistical Analysis

SPSS 22.0 software was used for statistical analysis. The data were tested for normality(One-sample kolmogorov-smirnov test, KS test). We present continuous measurements as mean (SD) if they are normally distributed or median (IQR) if they are not, and categorical variables as count (%).

### Result

Investigations into the sources of exposure indicate a history of contact with person from Hubei or family clusters in each case. There were 9 males (35%) and 17 females (65%), with an average age of 6.9 (0.7) years, ranging from 1 to 13 years (table 1). None of the above children had underlying diseases such as congenital diseases, dysplasia, diabetes, etc. Because some children cannot expresss independently, the clinical symptoms were mainly objective symptoms. Most patients had fever and cough at admission. About one-third of the patients had no clear symptoms, and other symptoms included vomiting, diarrhea, and rhinorrhoea (table 1). According to the standard,8 cases(31%) were mild and 18(69%) were ordinary. No severe complications such as acute respiratory distress syndrome (ARDS) and acute lung injury (ALI) occurred in the cases. Laboratory results revealed that most of the blood routine showed normal or decreased white blood cell counts, lymphocytes generally increased, and hemoglobin and platelets were mostly unaffected (table 2). There were no significant abnormalities in coagulation function. In terms of biochemistry, Alanine transaminase (ALT) combine with glutamic oxaloacetic transaminase (AST) increased in two cases(8%),ALT or AST increased in one case(4%).In addition, all the above cases had not exceed 50% of the normal high value. Most of the myocardial enzymes(myoglobin, troponin, kinase, creatine kinase) were not abnormal, with only a significant increase in LDH(46%). In the field of inflammatory mediators (table 2), procalcitonin was normal, and 5(19%) patients had elevated C-reative protein. 19 patients were tested for interleukin-6 and only one had an increase. T-lymphocyte subset analysis showed no abnormalities in CD3+CD4+/CD3+CD8+.Humoral immunity showed that IgA, IgG, and C3c were partially reduced (table 2), and IgM and C4 were normal. According to chest X-ray and chest CT examination, 8 patients(31%) had no pulmonary infiltration. There were 11 cases(42%) of lateral pulmonary infiltration and 7 cases(27%) of bilateral pulmonary infiltration.

**Table 1.** Baseline characteristics of the 26 children with 2019-nCoV

|  | Patients (n=26) |
| --- | --- |
| Age, years |  |
| Mean (SD) | 6.9 (0.7) |
| Range | 1-13 |
| 1-3 | 5 (19%) |
| 3-7 | 8 (31%) |
| 7-14 | 13 (50%) |
| Sex |  |
| Female | 9 (35%) |
| Male | 17 (65%) |
| Symptoms |  |
| No | 9 (35%) |
| Fever | 11 (42%) |
| Cough | 12 (46%) |
| Rhinorrhoea | 2 (8%) |
| Vomiting | 2 (8%) |
| Diarrhea | 2 (8%) |
| More than one symptoms | 10 (38%) |
| Discharged | 17 (65%) |
| Remain in hospital | 9 (35%) |

**Table 2.** Laboratory characteristics of the 26 children with 2019-nCoV

| Laboratory characteristics | Patients (n=26) |
| --- | --- |
| Leucocytes* |  |
| increase | 4 (15%) |
| decrease | 13 (50%) |
| Lymphocytes (normal range: $0.2-0.4 \times 10^9/L$) | |
| increase | 25 (96%) |
| decrease | 1 (4%) |
| Platelets (normal range: 100-300g/L) |  |
| normal | 18 (69%) |
| ALT>45U/L | 3 (12%) |
| AST>45/L | 3 (12%) |
| LDH>250U/L | 12 (46%) |
| ESR>15mmol/L | 7 (27%) |
| CRP>5.0mg/L | 5 (19%) |
| IgG<7 | 5 (19%) |
| IgA<0.7 | 6 (23%) |
| C3c | 4 (15%) |

\*<7 years old normal range: $8.0-10.0 \times 10^9/L$ ; 7-12years old normal
range: $4.5-13.5 \times 10^9/L$ ; >12years old normal range: $3.5-9.5 \times 10^9/L$

All patients were treated in isolation. Medicines for treating COVID-19 include oseltamivir, ribavirin, interferon, kaletra and traditional Chinese medicine.4 patients (15%) were treated with interferon alone and 1 patient (4%) was treated with kaletra alone.26 patients discharge (13.6±1.03 days on average) or improvement after treatment.

## DISCUSSION

There was no significant increase in inflammatory indexes and mild clinical symptoms in 26 patients, all of them were mild and common type cases, no severe pneumonia cases and death cases, and the outcome was similar to that of SARS. Among them, 11 patients (about 42%) had fever, which was inconsistent with 98% of the clinical symptoms of adult patients reported in [2,3]. Other symptoms such as nasal obstruction, runny nose, vomiting and diarrhea [1,3,6] were less common than adults. All of the above suggest that the clinical symptoms of children infected with 2019-CoV were not typical. It is considered that most of the children might be infected after exposure to patients with COVID-19, which belongs to the second or third generation of infected cases, and the pathogenicity of the virus is decreased.

In addition, the changes of leukocytes in children were the same as in adults, and most of them appear normal or decreased. Lymphocytes in adult patients had decreased significantly, and this study found that most of the children’s lymphocytes increased beyond the normal range, which might be related to the higher percentage of lymphocytes in white blood cells in the process of children’s growth and development, and then gradually decreased towards adults. This might also indicate that 2019-ncov produces a series of immune responses in the body through the respiratory mucosa, and children’s lymphocytes had not significantly decreased, thus failing to induce a cytokine storm. Chen N et al.[3] studied the imaging characteristics of 99 adult 2019-CoV pneumonia patients in Wuhan area and found that 75% of patients had double pneumonia, and the remaining 25% of patients had single pneumonia. We found that 8 of the 26 children did not have pneumonia, 7 (about 27%) had bilateral pulmonary infiltration, and 11 (about 42%) had unilateral pulmonary infiltration. It can be seen that the CT manifestations of children are quite different from that of adults, and complications such as ARDS have not been found in children in China so far, which can indirectly reflect that children are not easy to induce not prone to induce cytokine storms. The literature reported that the prognosis of elderly patients and those with chronic underlying diseases is poor, which was presumed to be associated with low immunity [3]. We know that most T cells develop in the thymus. In this study, the prognosis of children with COVID-19 was good, and the prognosis was different from that of middle-aged and elderly patients in other studies. Consider that the difference was related to thymic immune function. In the treatment of SARS patients, drugs such as glucocorticoids and thymosin were used [7]. Although there was no clinical evidence to support the use of glucocorticoids [8], whether early use of thymosin and gamma globulin for immunomodulation in patients with severe and critical COVID-19 can reduce cytokine storms, reduce clinical symptoms and improve prognosis requires further exploration.

Except that LDH increased in the myocardial zymogram of children with COVID-19 in the study, other enzymes such as creatine kinase and troponin I were normal. In addition to being derived from the heart muscle, LDH was also found in the liver and kidneys. We speculated that the increase in LDH may not originate from the myocardium, suggesting that children’s infections were less likely to affect the myocardium. Similar to other studies [1, 3, 4], this study found that ALT and AST increased, so we cannot rule out the effect of 2019-nCoV on the liver and induce the increase of liver enzymes. As patients often use antiviral drugs in the early stage of hospitalization, it was also necessary to consider the increase of liver enzymes caused by drug factors. Angiotensin converting enzyme 2 (ACE2) had been shown to be one of the main receptors that mediate the entry of 2019-nCoV into human cells [9, 10], while ACE2 was at high expression levels in the kidney, vas deferens and testicular mesenchymal cells[11]. SARS had been shown to cause orchitis and affect spermatogenesis function [11, 12].Although acute renal impairment had not yet occurred in pediatric cases, children with COVID-19 must be alert to potential risks and follow-up on reproductive function after discharge from hospital.

Patients in this study were tested with the 2019-nCOV nucleic acid test. The specimens included nasopharyngeal swabs, blood, sputum, and anal swabs. We found that nasal swabs and anal swabs were positive even without respiratory symptoms such as cough and gastrointestinal symptoms such as vomiting and diarrhea Therefore, the source of samples cannot be selected based on clinical manifestations, and should be submitted multiple times and at multiple sites to avoid misdiagnosis of patients with mild imaging without imaging manifestations. Although there is no clinical evidence that the virus is transmitted through the conjunctival pathway [13], mother-infant, and breast-milk[4], researchers have detected the virus in urine [14], saliva [15], suggesting that novel coronavirus has a wide range of transmission routes.

The clinical classification of children was mostly mild or ordinary type, so the treatment plan is relatively simple. In some cases, the nucleic acid test turned negative after treatment with interferon atomization or lopinavir ritonavir. For mild children, symptomatic treatment or single use of traditional Chinese medicine needs to be further verified.

The sample size of this study was only 26, and it was a retrospective study, so it has certain limitations. On the one hand, the article only included children aged 1-14 years. The characteristics of newborns infected with COVID-19 are not clear, and whether there are differences in the disease characteristics of infants, preschool and school-age children need further research; On the other hand, the changes of T lymphocyte subsets in children need to be tracked and observed during the course of the disease.

In summary, COVID-19 children showed asymptomatic or mainly respiratory symptoms such as fever and dry cough, some of them have digestive tract symptoms, and their clinical symptoms are mild. In our study, the clinical types were mild and common, and the probability of severe complications such as myocarditis and ARDS was low, which provided important information for understanding the clinical characteristics of children with COVID-19.

## Data Availability

All data is available.All data used during the study appear in the submitted article.

## DATA AVAILABILITY STATEMENT

All data used during the study appear in the submitted article.

## ACKNOWLEDGMENTS

The authors thank the department of Shenzhen Center for Disease Control and Prevention(CDC) and the hepatopathy institute of National Infectious Disease Clinical Medical Research Center (The Third People’s Hospital of Shenzhen) for the 2019-nCoV nucleic acid tests.

## Notes

### Competing Interest Statement

The authors have declared no competing interest.

### Funding Statement

This work was supported by The Project of Sanming
Foundation of Shenzhen, China (Item coding: SZSM201911009, SZSM201612025)

